# Walking Activities during the Acute Stroke Hospital Stay May Benefit Cerebrovascular Health

**DOI:** 10.1101/2021.06.10.21258640

**Authors:** Alicen A. Whitaker, Madison L. Henry, Allegra Morton, Jaimie L. Ward, Sarah M. Eickmeyer, Michael G. Abraham, Sandra A. Billinger

**Affiliations:** Department of Physical Therapy and Rehabilitation Science, University of Kansas Medical Center, Kansas City, KS; Department of Physical Medicine and Rehabilitation, University of Kansas Medical Center, Kansas City, KS; Department of Neurology, University of Kansas Medical Center, Kansas City, KS; Department of Integrative and Molecular Physiology, University of Kansas Medical Center, Kansas City, KS; Department of Interventional Radiology, University of Kansas Medical Center, Kansas City, KS

**Keywords:** Cardiovascular Rehabilitation, Middle Cerebral Artery Blood Velocity, Brain Health

## Abstract

**Purpose:** Physical activity within the hospital post-stroke is recommended for cardiovascular and musculoskeletal health, but no studies have examined cerebrovascular health. We hypothesized individuals who walked farther distances (FARhigh) in the hospital would have a higher resting middle cerebral artery blood velocity (MCAv) and a greater cerebrovascular response (CVR) to moderate-intensity exercise at 3-months post-stroke, compared to individuals who walked shorter distances (FARlow).

**Methods:** At 3-month post-stroke, we recorded 90-seconds of baseline (BL) MCAv followed by 6-minutes of moderate-intensity exercise. We calculated CVR as the change in MCAv from BL to steady-state exercise. We collected farthest distance walked from the electronic medical record. We divided individuals based on average farthest walking distance, (FARhigh or FARlow).

**Results:** Participants (n = 20) were 63 ± 15 years. BL MCAv was not different between groups (p = 0.07). In comparison to FARlow, we report a higher CVR in FARhigh’s ipsilesional (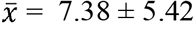 vs 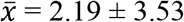, p = 0.02) and contralesional hemisphere (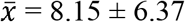 vs 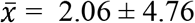, p = 0.04).

**Conclusions:** Physical activity during the hospital stay post-stroke may support cerebrovascular health after discharge. Prospective studies are needed to support this finding.

## Introduction and Purpose

After stroke, light to moderate physical activity is recommended during the acute hospital stay to improve cardiovascular and musculoskeletal health.^1^ Physical activity is also recommended for cerebrovascular health in aging older adults and individuals with stroke.^2-5^ However, it is well documented that individuals post-stroke within the acute hospital spend the majority of their day inactive.^6-9^ Recently, research initiatives are investigating increased physical activity during the acute hospital stay post-stroke.^1^ While studies have examined motor function and self-reported disability outcomes with increased physical activity during the acute hospital stay,^10-12^ no studies have examined the cerebrovascular system. Adequate perfusion and regulation of cerebral blood flow is essential for the brain to function properly. Therefore, stroke rehabilitation research should include measures of cerebrovascular health to increase our understanding regarding the relationship between physical activity and brain health.^13^

Our prior work has shown middle cerebral artery blood velocity (MCAv), a surrogate measure of cerebral blood flow, is constant at rest and increases with submaximal exercise in healthy aging adults and stroke.^14-18^ We report no significant differences in MCAv during seated rest in individuals at 3-months post-stroke compared to age- and sex-matched adults.^18^ However, when we challenged the system with moderate-intensity exercise, individuals 3-months post-stroke had a reduced cerebrovascular response (CVR) compared to controls.^18^ CVR is defined as the change in MCAv from rest to steady-state exercise and has been positively associated with cardiovascular health in older adults^19^ and aerobic fitness in individuals with stroke.^17^ Further investigation in individuals at 3- and 6-months post-stroke showed a greater CVR to moderate-intensity exercise in individuals who reported being physically active and had a higher non-exercise estimated maximal oxygen uptake (VO_2max_) compared to those who reported being inactive or sedentary.^17^ Collectively, these findings suggest physical activity engagement could prove beneficial to cerebrovascular health following stroke. Therefore, the next step in our line of research was to explore physical activity within the acute hospital and cerebrovascular health post-stroke.

The objective of this secondary analysis was to assess walking during the acute stroke hospital stay and MCAv at 3-months. We hypothesized that individuals who walked farther within the acute hospital post-stroke (FARhigh) would have 1) a higher resting MCAv at baseline (BL) and 2) a greater CVR at 3-months post-stroke, compared to individuals who walked shorter distances (FARlow).

## Methods

### Recruitment

The primary study, characterizing the longitudinal changes in MCAv at rest and during exercise post-stroke, has previously published the recruitment^20^ and methodology details.^17^ Briefly, participants were included if they met the following criteria: 1) Age 35 to 95 years old, 2) unilateral ischemic stroke, 3) less than 70% carotid stenosis observed on imaging, 4) physician approval to perform moderate-intensity exercise, and 5) able to walk more than 10 meters without physical assistance. Participants were excluded if they 1) were unable to consent due to global aphasia, 2) were not able to perform exercise on a seated stepper, 3) had another neurologic disorder such as Parkinson’s disease, Alzheimer’s disease, or Multiple Sclerosis, or 4) were dependent on supplemental oxygen. For this secondary analysis, we also excluded participants if their acute hospital stay was not at the University of Kansas Health System, where the walking data was collected from the electronic medical record (EMR). The University of Kansas Medical Center Human Subjects Committee approved this study and all the experimental procedures. Participants provided informed written consent prior to the start of study procedures.

### Electronic Medical Record

We used the University of Kansas Health System’s daily activity spreadsheet within the EMR to examine walking during the acute hospital stay post-stroke. The interdisciplinary teams at the University of Kansas Hospital System monitors patient progress through the daily activity spreadsheet, where walking distance (in feet) and frequency of out-of-bed activities are recorded. The walking data collected from the daily activity spreadsheet was standard of care on the stroke unit. No information was available whether patients with stroke were following published early out of bed activity or rigorous walking protocols.^10,21^

### Study Visit at 3-Months Post-Stroke

Prior to the visit, participants were asked to refrain from caffeine for 6 hours, food for 2 hours, and vigorous exercise for 12 hours. Participants came to the laboratory at 3-months post-stroke for the study visit. As previously published, we collected demographics and past medical information, competed questionnaires, and performed an exercise familiarization session prior to the MCAv recordings.^17^ Participants practiced stepping on a recumbent stepper (T5XR NuStep, Inc. Ann Arbor, MI) at a constant rate of 90 steps per minute while we determined the resistance for moderate-intensity exercise (45-55% heart rate reserve). ^22^ If a participant was on beta-blocker medication the estimated maximal heart rate equation was used (164 – (0.7 x age)).^23^ To ensure accurate measures of end-tidal carbon dioxide (P_ET_CO_2_), participants also practiced breathing through their nose during exercise.

#### MCAv Recording

Next, we donned the following equipment: 1) 2-MHz transcranial Doppler ultrasound (TCD) probes secured with an adjustable headband (Multigon Industries Inc, Yonkers, New York) to monitor bilateral MCAv, 2) a 5-lead electrocardiogram ((ECG; Cardiocard, Nasiff Associates, Central Square, New York) to monitor HR, 3) a nasal cannula attached to a capnography (BCI Capnocheck Sleep 9004 Smiths Medical, Dublin, Ohio) to monitor P_ET_CO_2_, and 4) a beat-to-beat blood pressure cuff on the left middle finger (Finometer, Finapres Medical Systems, Amsterdam, the Netherlands) to monitor mean arterial pressure (MAP).

Aligned with our prior work, we calculated BL MCAv as an average of the 90 second seated rest before the onset of moderate-intensity exercise. CVR was calculated as the difference between BL and the average steady state exercise MCAv (minutes 3 to 4.5).^14,17,18,24^ After the cool down, participants rested to allow HR to return to near BL values. Following the rest, participants performed a 6-minute walk test (6MWT) in a hallway with no distractions. We conducted the 6MWT according to the guidelines from the American Thoracic Society.^25^

### Statistical Analysis

Data are reported as means ± standard deviations unless otherwise indicated. Alpha was set a priori at ≤ 0.05. Normality for each variable was determined using a Shapiro-Wilk test. Based on the farthest walking data collected from the EMR, participants were divided into groups.

Individuals that walked more than the average farthest distance (FARhigh) during the acute hospital stay were compared to individuals who walked less than the average farthest distance (FARlow). Participant characteristics were compared between FARhigh and FARlow using a One-way Analysis of Variance (ANOVA), Kruskal-Wallis test for non-parametric variables, and Fisher exact test for categorical variables. Measures of MCAv and exercise fidelity were compared between groups using a One-way Analysis of Variance (ANOVA) and Kruskal-Wallis test for non-parametric variables.

## Results

Of the 27 participants consented into the primary study, 20 individuals were included within this secondary analysis (40% women, age 63 ± 15 years).^17^ One participant had 2 stroke events separated by one year. Due to our interest in comparing the ipsilesional and contralesional MCAv, the second stroke was excluded within this analysis.^24^ We also excluded individuals who did not have a valid MCAv signal at rest (n=3) or during exercise (n=1) and individuals who were not admitted to the University of Kansas Health System (n=2) as we were interested in the daily activity tracker within their electronic medical record. There were no significant differences between groups for stroke characteristics or treatment interventions, shown in **Table 1**.

**Table 1.** Participant Characteristics.

|  | *FARhigh (n=11)* | *FARlow (n=9)* | *P value* |
| --- | --- | --- | --- |
| **Collected at 3-Months** |  |  |  |
| Age (years) | 59 (16) | 68 (14) | 0.18 |
| Females (n (%)) | 4 (36%) | 4 (44%) | 1.00 |
| Race (n (%)) * |  |  |  |
| African American | 3 (27%) | 4 (44%) | 0.64 |
| Native American | 1 (9%) | 0 | 1.00 |
| Asian | 1 (9%) | 0 | 1.00 |
| Caucasian | 8 (73%) | 5 (56%) | 0.64 |
| Body Mass Index | 30.7 (5.2) | 29.1 (6.3) | 0.55 |
| Beta Blocker Medication (n (%)) | 7 (64%) | 9 (100%) | 0.09 |
| Modified Rankin Scale (n (%)) |  |  |  |
| No Symptoms | 0 | 0 |  |
| No Significant Disability | 4 (36%) | 2 (22%) |  |
| Slight Disability | 4 (36%) | 5 (56%) |  |
| Moderate Disability | 2 (18%) | 1 (11%) |  |
| Moderately Severe Disability | 1 (9%) | 1 (11%) |  |
| Severe Disability | 0 | 0 | 0.91 |
| Six Minute Walk Test (m) | 340.3 (103.5) | 264.6 (160.3) | 0.22 |
| **Collected from EMR** |  |  |  |
| Number of Days Between Stroke<br>and Study Visit | 93 (10) | 99 (15) | 0.29 |
| Length of Hospital stay (days) | 6 (2.8) | 5 (1.8) | 0.50 |
| Lesion Size (cm<sup>3</sup>) ^ | 71.9 (99.2) | 6.7 (10.4) | 0.27 |
| NIH Stroke Scale upon Admission† | 10 (7.6) | 6 (5.1) | 0.13 |
| TPA (n (%)) | 5 (45%) | 3 (33%) | 0.67 |
| Thrombectomy (n (%)) | 4 (36%) | 1 (11%) | 0.32 |

Mean (Standard Deviation) unless otherwise indicated. \*One participant selected multiple races. EMR = Electronic Medical Record. NIH = National Institutes of Health. TPA = Tissue Plasminogen Activator. ^ = FARhigh (n=10) and FARlow (n=7). † = FARlow (n=8)

The participants’ average farthest walking distance within the acute hospital post-stroke was 340 feet (SD = 225). FARhigh, characterized as ≥ 340 feet, had an average farthest walking distance of 508 feet (SD = 136). FARlow, characterized as < 340 feet, had an average farthest walking distance of 134 feet (SD = 103). We also examined the frequency these individuals walked per day during the acute hospital stay and found no significant difference between FARhigh (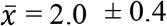 times per day,) and FARlow (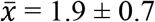 times per day, p = 0.53). At the visit 3-months post-stroke, the 6MWT distance, revealed no significant difference between group differences (p = 0.22). However, we report the difference was clinically meaningful with FARhigh walking 75 meters farther than FARlow.^26^

### MCAv at 3-Months Post-Stroke

We found no significant differences between FARhigh and FARlow for BL MCAv within the ipsilesional and contralesional hemisphere, shown in **Table 2**. One individual within the FARlow group had a high contralesional resting MCAv. We assessed whether the data met criteria for an outlier and warranted removal from the data analysis. The resting MCAv data point was determined to not be an outlier and remained for all analyses. During moderate-intensity exercise, FARhigh had a greater CVR within the ipsilesional hemisphere compared to FARlow (p = 0.02). The CVR to moderate-intensity exercise within the contralesional hemisphere was also greater within FARhigh compared to FARlow (p = 0.04).

**Table 2.** MCAv Response between FARhigh and FARlow.

|  |  | *FAR<sub>high</sub>*<br>(*n*=11) | *FAR<sub>high</sub>*<br>95% *CI* | *FAR<sub>low</sub>*<br>(*n*=9) | *FAR<sub>low</sub>*<br>95% *CI* | *P* value |
| --- | --- | --- | --- | --- | --- | --- |
| **Ipsilesional Hemisphere** |  |  |  |  |  |  |
|  | BL MCAv<br>(cm/s) | 46.83 (11.32) | 40.16 – 53.37 | 59.99 (18.82) | 46.80 – 71.90 | 0.07 |
|  | CVR<br>(cm/s) | 7.38 (5.42) | 4.35 – 10.73 | 2.19 (3.53) | 0.02 – 4.40 | **0.02** |
| **Contralesional Hemisphere\*** |  |  |  |  |  |  |
|  | BL MCAv<br>(cm/s) | 54.36 (10.37) | 47.85 – 60.79 | 75.09 (30.57) | 56.37 – 99.30 | 0.07 |
|  | CVR<br>(cm/s) | 8.15 (6.37) | 4.47 – 11.96 | 2.06 (4.76) | -1.42 – 5.31 | **0.04** |

Mean (Standard Deviation). 95% Confidence Intervals. BL = Baseline, MCAv = Middle Cerebral Artery Blood Velocity, cm/s = centimeters per second, CVR = Cerebrovascular Response. \*FAR<sub>high</sub> (n=10) and FAR<sub>low</sub> (n=8)

### Blood Pressure, Respiration, and Heart Rate

MAP, P_ET_CO_2_, and HR at BL and steady state exercise are shown in **Table 3**. MAP and HR were not different between groups and were unlikely to contribute to the difference in CVR between groups. While P_ET_CO_2_ was significantly higher in the FARhigh group, which could result in a higher MCAv, the change from BL to steady state exercise P_ET_CO_2_ was not different between FARhigh 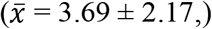 and FARlow 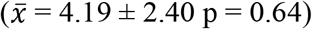. Therefore, the change in P_ET_CO_2_ was not likely the driving factor for a higher CVR within the FARhigh group. Both groups also performed similar effort during the moderate-intensity exercise as workload, HR, and RPE were not different.

**Table 3.**
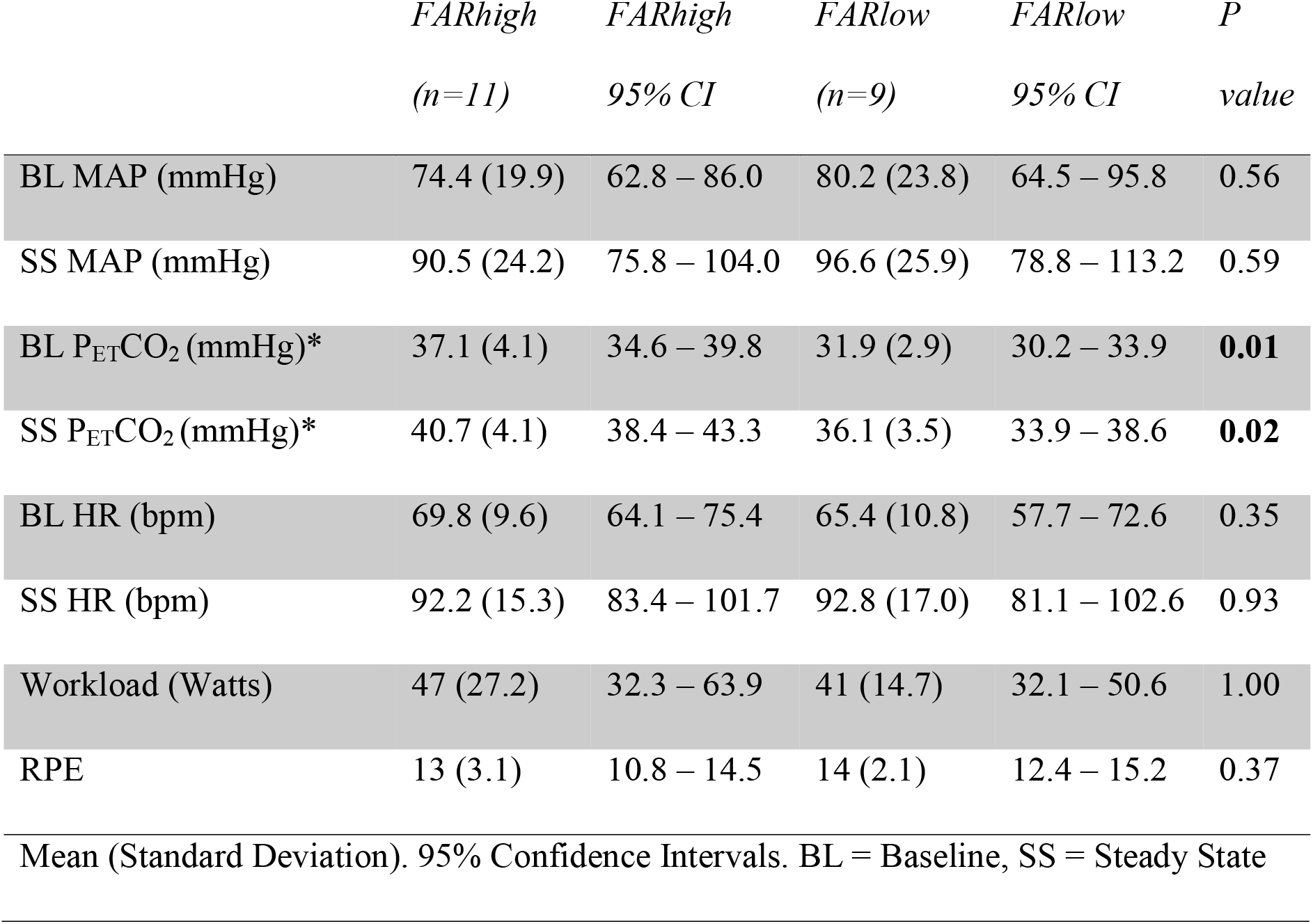
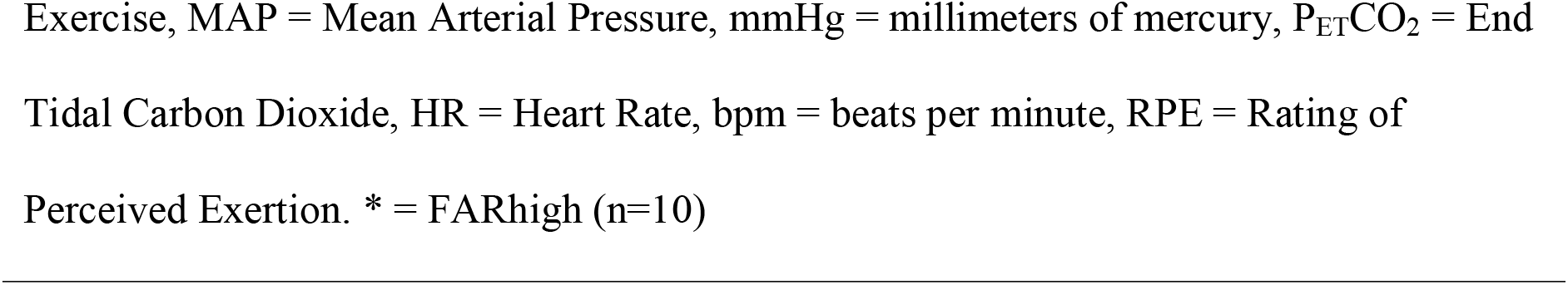
Exercise Fidelity Measures.

|  | *FARhigh*<br>(*n*=11) | *FARhigh*<br>95% CI | *FARlow*<br>(*n*=9) | *FARlow*<br>95% CI | *P*<br>value |
| --- | --- | --- | --- | --- | --- |
| BL MAP (mmHg) | 74.4 (19.9) | 62.8 – 86.0 | 80.2 (23.8) | 64.5 – 95.8 | 0.56 |
| SS MAP (mmHg) | 90.5 (24.2) | 75.8 – 104.0 | 96.6 (25.9) | 78.8 – 113.2 | 0.59 |
| BL $P_{ET}CO_2$ (mmHg)* | 37.1 (4.1) | 34.6 – 39.8 | 31.9 (2.9) | 30.2 – 33.9 | **0.01** |
| SS $P_{ET}CO_2$ (mmHg)* | 40.7 (4.1) | 38.4 – 43.3 | 36.1 (3.5) | 33.9 – 38.6 | **0.02** |
| BL HR (bpm) | 69.8 (9.6) | 64.1 – 75.4 | 65.4 (10.8) | 57.7 – 72.6 | 0.35 |
| SS HR (bpm) | 92.2 (15.3) | 83.4 – 101.7 | 92.8 (17.0) | 81.1 – 102.6 | 0.93 |
| Workload (Watts) | 47 (27.2) | 32.3 – 63.9 | 41 (14.7) | 32.1 – 50.6 | 1.00 |
| RPE | 13 (3.1) | 10.8 – 14.5 | 14 (2.1) | 12.4 – 15.2 | 0.37 |

Mean (Standard Deviation). 95% Confidence Intervals. BL = Baseline, SS = Steady State
Exercise, MAP = Mean Arterial Pressure, mmHg = millimeters of mercury, $P_{ET}CO_2$ = End Tidal Carbon Dioxide, HR = Heart Rate, bpm = beats per minute, RPE = Rating of Perceived Exertion. \* = FARhigh (n=10)

## Discussion

With recent research, such as the A Very Early Rehabilitation (AVERT) trial, underlying the importance of dosing physical activity within the acute hospital for stroke recovery at 3-months, this study contributes toward filling the gaps in knowledge. We hypothesized that individuals who performed more physical activity, such as walking father, during the acute hospital stay post-stroke would have a higher resting MCAv and greater CVR to moderate-intensity exercise at 3-months. While our hypotheses were only partially supported by our findings, those who walked farther distances within the hospital had a greater CVR to moderate-intensity exercise at 3-months. Therefore, we report standard of care walking and motivating individuals to perform physical activity during the acute hospital stay post-stroke may contribute to cerebrovascular health with recovery.

### Resting MCAv

Our study found no significant differences in BL MCAv between FARhigh and FARlow. Our finding is similar to previous work showing no differences in resting MCAv with more out-of-bed mobility at 52 hours post-stroke compared to less out-of-bed mobility at 7 days.^27^ This result on the contralesional MCAv may be driven by the individual with the higher resting value. As shown in Table 2, the FARlow group shows a large standard deviation and is likely influencing these results. We acknowledge the small sample size may be a contributing factor. Despite these limitations, the findings contribute novel information and begin to address a scientific gap in knowledge. We acknowledge many factors may influence the resting MCAv values 3 months later such as medication, spontaneous stroke recovery, participation in physical therapy interventions and physical activity engagement after discharge.

### Cerebrovascular Response

Individuals who walked farther distances within the acute hospital had a higher CVR to the exercise stimuli at 3-months post-stroke. While blood pressure, heart rate, and carbon dioxide increased from baseline to moderate-intensity exercise, the change scores were not significantly different between groups and were unlikely to contribute to the difference in CVR. Instead, the higher CVR to an acute bout of exercise could be due to the positive influence of physical activity and walking on vascular health by reducing oxidative stress and increasing vasomotor reactivity^28-31^ during the hospital stay. Previous work reported an 8-week physical activity intervention during subacute stroke reduced oxidative stress^32^ and a 6-month walking intervention in chronic stroke increased the ability of the cerebrovascular system to vasodilate and vasoconstrict.^31^ Our previous work has shown that self-reported physical activity at 3- and 6-months post stroke was associated with a greater CVR than those reporting low physical activity levels.^17^ Indeed, increased physical activity within the acute hospital and after discharge may contribute to a greater responsiveness of the cerebrovascular system during stroke recovery. Future work should study physical activity through accelerometry or mobile devices at hospital discharge to ensure better physical activity characterization. This information would increase our understanding regarding how physical activity influences cerebrovascular health following stroke. Our findings, presented here, provide foundational information regarding out of bed activity during the hospital stay and cerebrovascular health at 3 months post-stroke.

### Walking Frequency and Endurance

The frequency walked during the acute hospital stay was standard of care and did not follow the recommendations from the AVERT trial. The AVERT trial reported individuals who performed short bouts of physical activity approximately 10x/day had more favorable outcomes at 3-months post-stroke, such as lower self-reported disability symptoms.^13,21^ Therefore, an average walking frequency of 2x/day, reported within this study, may be too low to have an impact on cerebrovascular health. Previous studies in healthy adults have also reported a frequency of walking every 30 minutes, preserves resting MCAv throughout the day compared to sedentary prolonged sitting.^33,34^ Therefore, walking at a greater frequency may be needed to increase resting MCAv in individuals post-stroke.^10,33,34^

While this study separated individuals into groups based on the average farthest distance walked during the acute stroke hospital stay, functionally, 340 feet is the ability of an individual to walk the length of a soccer field or low-level walking within the community.^35^ Therefore, further walking during the acute hospital stay could have encouraged individuals to continue performing physical activity within the community after discharge from the hospital. While not statistically significant, this study did show that individuals who walked farther during the acute hospital stay also had a clinically meaningful higher walking endurance during the 6MWT at 3-months. The study’s higher walking endurance of more than 50 meters during the 6MWT has been reported to be “substantially meaningful” to quality of life.^26^ Although we didn’t collect information regarding lower extremity function or balance, the modified Rankin score at our 3 months post-stroke visit suggests no significant differences for disability between the groups. Therefore, lower extremity function may not be a primary factor for the 6MWT distance and during the acute exercise bout.

### Role of Physical Therapy for Cerebrovascular Health

This study is timely for physical therapy, as activity within the hospital has been highlighted as an important factor for stroke recovery that requires further research.^1,10,13^ Physical therapists and physical therapy assistants have instrumental roles in advocating for and providing out-of-bed activities that may contribute to long-term cerebrovascular health. Recent reviews underscore the importance of mobility interventions post-stroke (i.e. head-of-bed tilt and aerobic exercise) for MCAv, which could be significant for rehabilitation success.^13,36^ After a stroke, one study reported a higher MCAv upon admission to inpatient stroke rehabilitation was associated with improved Functional Independence Measure (FIM) scores at discharge.^37^ Studies in healthy adults also report prolonged inactivity was associated with a decrease in MCAv,^33,38^ cardiovascular mortality,^39^ and a longer cognitive reaction time.^40^ Further research is needed to determine whether performing physical activity and walking farther distances within the acute hospital to increase measures of MCAv is associated with improved functional recovery during inpatient rehabilitation.

### Limitations

While this study provides an important future direction for prospectively collecting physical activity during the acute hospital and cerebrovascular health during stroke recovery, there are several limitations to consider due to the retrospective study design. First, walking data during the acute hospital stay was not prospectively collected and we were also not able to collect information regarding the length of time from stroke onset to the start of walking within the hospital. Therefore, we are unable to provide further details on the timeline to begin walking further distances post-stroke. Second, heart rate was not recorded during walking within the acute hospital stay and therefore we do not know what intensity these individuals were walking. Third, this study was not powered to detect statistical significance. Due to a small sample size, confounding factors such as age, sex, and medications were not statistically controlled. Finally, MCAv is linearly related to blood flow when a constant MCA diameter is maintained,^41^ which has been shown during various conditions including moderate-intensity exercise.^42-44^ Consistent with our previously published work in stroke, we assumed arterial diameter was maintained.^17,18^

## Conclusions

This retrospective analysis is the first to report increased physical activity during the acute hospital stay and a greater cerebrovascular response at 3-months post-stroke. This study suggests that physical therapists may impact motor function as well as brain health by providing the motivation, support, and patient education needed to be more physically active during the acute hospital stay post-stroke. Physical therapists need to consider physical activity for cerebrovascular health post-stroke as it relates to cognition,^45^ neuroplasticity,^46^ and functional outcomes.^37,47^ A prospective research study to confirm these retrospective findings is the next necessary step for neurovascular stroke rehabilitation.

## Supporting information

COI Disclosure

COI Disclosure

COI Disclosure

STROBE Checklist

COI Disclosure

COI Disclosure

COI Disclosure

COI Disclosure

## Data Availability

Data is available upon request to corresponding author.

## Acknowledgements

Dr. Billinger was supported by the Eunice Kennedy Shriver National Institute of Child Health and Human Development (K01HD067318). Dr. Whitaker, Ms. Morton, and Ms. Henry received support by the Eunice Kennedy Shriver National Institute of Child Health and Human Development (T32HD057850). Ms. Morton received a Student Scholarship in Cerebrovascular Disease and Stroke from the American Heart Association. REDCap at KU Medical Center is supported by the Clinical and Translational Science Award (UL1TR000001) from the National Center for Research Resources and National Center for Advancing Translational Science. The National Center for Advancing Translational Science awarded to the University of Kansas Frontiers the Clinical and Translational Science Institute (UL1TR002366). The contents are solely the responsibility of the authors and do not necessarily represent the official views of the NIH.

## References

1. Billinger SA, Arena R, Bernhardt J, et al. Physical activity and exercise recommendations for stroke survivors: a statement for healthcare professionals from the American Heart Association/American Stroke Association. Stroke. 2014;45(8):2532–2553.

2. Izzo C, Carrizzo A, Alfano A, et al. The Impact of Aging on Cardio and Cerebrovascular Diseases. Int J Mol Sci. 2018;19(2).

3. Benedictus MR, Leeuwis AE, Binnewijzend MA, et al. Lower cerebral blood flow is associated with faster cognitive decline in Alzheimer’s disease. Eur Radiol. 2017;27(3):1169–1175.

4. Bangen KJ, Werhane ML, Weigand AJ, et al. Reduced Regional Cerebral Blood Flow Relates to Poorer Cognition in Older Adults With Type 2 Diabetes. Front Aging Neurosci. 2018;10:270.

5. Schuff N, Matsumoto S, Kmiecik J, et al. Cerebral blood flow in ischemic vascular dementia and Alzheimer’s disease, measured by arterial spin-labeling magnetic resonance imaging. Alzheimers Dement. 2009;5(6):454–462.

6. Mattlage AE, Redlin SA, Rippee MA, Abraham MG, Rymer MM, Billinger SA. Use of Accelerometers to Examine Sedentary Time on an Acute Stroke Unit. J Neurol Phys Ther. 2015;39(3):166–171.

7. Bernhardt J, Dewey H, Thrift A, Donnan G. Inactive and alone: physical activity within the first 14 days of acute stroke unit care. Stroke. 2004;35(4):1005–1009.

8. Sjoholm A, Skarin M, Churilov L, Nilsson M, Bernhardt J, Linden T. Sedentary behaviour and physical activity of people with stroke in rehabilitation hospitals. Stroke Res Treat. 2014;2014:591897.

9. Astrand A, Saxin C, Sjoholm A, et al. Poststroke Physical Activity Levels No Higher in Rehabilitation than in the Acute Hospital. J Stroke Cerebrovasc Dis. 2016;25(4):938–945.

10. Langhorne P, Wu O, Rodgers H, Ashburn A, Bernhardt J. A Very Early Rehabilitation Trial after stroke (AVERT): a Phase III, multicentre, randomised controlled trial. Health Technol Assess. 2017;21(54):1–120.

11. Sundseth A, Thommessen B, Ronning OM. Outcome after mobilization within 24 hours of acute stroke: a randomized controlled trial. Stroke. 2012;43(9):2389–2394.

12. Liu N, Cadilhac DA, Andrew NE, et al. Randomized controlled trial of early rehabilitation after intracerebral hemorrhage stroke: difference in outcomes within 6 months of stroke. Stroke. 2014;45(12):3502–3507.

13. Marzolini S, Robertson AD, Oh P, et al. Aerobic Training and Mobilization Early Post-stroke: Cautions and Considerations. Front Neurol. 2019;10:1187.

14. Billinger SA, Craig JC, Kwapiszeski SJ, et al. Dynamics of middle cerebral artery blood flow velocity during moderate-intensity exercise. J Appl Physiol (1985). 2017;122(5):1125–1133.

15. Ward JL, Craig JC, Liu Y, et al. Effect of healthy aging and sex on middle cerebral artery blood velocity dynamics during moderate-intensity exercise. Am J Physiol Heart Circ Physiol. 2018;315(3):H492–H501.

16. Witte E, Liu Y, Ward JL, et al. Exercise intensity and middle cerebral artery dynamics in humans. Respir Physiol Neurobiol. 2019;262:32–39.

17. Billinger SA, Whitaker AA, Morton A, et al. Pilot Study to Characterize Middle Cerebral Artery Dynamic Response to an Acute Bout of Moderate Intensity Exercise at 3- and 6-Months Poststroke. J Am Heart Assoc. 2021;10(3):e017821.

18. Kempf KS, Whitaker AA, Lui Y, et al. The Effect of Stroke on Middle Cerebral Artery Blood Flow Velocity Dynamics During Exercise. J Neurol Phys Ther. 2019;43(4):212–219.

19. Perdomo SJ, Ward J, Liu Y, et al. Cardiovascular disease risk is associated with middle cerebral artery blood flow velocity in older adults. Cardiopulm Phys Ther J. 2020;31(2):38–46.

20. Morton A, Myers M, Whitaker AA, et al. Optimizing Recruitment Strategies and Physician Engagement for Stroke Recovery Research. J Neurol Phys Ther. 2021;45(1):41–45.

21. Bernhardt J, Churilov L, Ellery F, et al. Prespecified dose-response analysis for A Very Early Rehabilitation Trial (AVERT). Neurology. 2016;86(23):2138–2145.

22. Camarda SR, Tebexreni AS, Pafaro CN, et al. Comparison of maximal heart rate using the prediction equations proposed by Karvonen and Tanaka. Arq Bras Cardiol. 2008;91(5):311–314.

23. Brawner CA, Ehrman JK, Schairer JR, Cao JJ, Keteyian SJ. Predicting maximum heart rate among patients with coronary heart disease receiving beta-adrenergic blockade therapy. Am Heart J. 2004;148(5):910–914.

24. Kaufman CS, Bai SX, Ward JL, Eickmeyer SM, Billinger SA. Middle cerebral artery velocity dynamic response profile during exercise is attenuated following multiple ischemic strokes: a case report. Physiol Rep. 2019;7(21):e14268.

25. Laboratories ATSCoPSfCPF. ATS statement: guidelines for the six-minute walk test. Am J Respir Crit Care Med. 2002;166(1):111–117.

26. Perera S, Mody SH, Woodman RC, Studenski SA. Meaningful change and responsiveness in common physical performance measures in older adults. J Am Geriatr Soc. 2006;54(5):743–749.

27. Diserens K, Moreira T, Hirt L, et al. Early mobilization out of bed after ischaemic stroke reduces severe complications but not cerebral blood flow: a randomized controlled pilot trial. Clin Rehabil. 2012;26(5):451–459.

28. Billinger SA, Mattlage AE, Ashenden AL, Lentz AA, Harter G, Rippee MA. Aerobic exercise in subacute stroke improves cardiovascular health and physical performance. J Neurol Phys Ther. 2012;36(4):159–165.

29. Palmefors H, DuttaRoy S, Rundqvist B, Borjesson M. The effect of physical activity or exercise on key biomarkers in atherosclerosis--a systematic review. Atherosclerosis. 2014;235(1):150–161.

30. Luca M, Luca A. Oxidative Stress-Related Endothelial Damage in Vascular Depression and Vascular Cognitive Impairment: Beneficial Effects of Aerobic Physical Exercise. Oxid Med Cell Longev. 2019;2019:8067045.

31. Ivey FM, Ryan AS, Hafer-Macko CE, Macko RF. Improved cerebral vasomotor reactivity after exercise training in hemiparetic stroke survivors. Stroke. 2011;42(7):1994–2000.

32. Taty Zau JF, Costa Zeferino R, Sandrine Mota N, et al. Exercise through a cardiac rehabilitation program attenuates oxidative stress in patients submitted to coronary artery bypass grafting. Redox Rep. 2018;23(1):94–99.

33. Carter SE, Draijer R, Holder SM, Brown L, Thijssen DHJ, Hopkins ND. Regular walking breaks prevent the decline in cerebral blood flow associated with prolonged sitting. J Appl Physiol (1985). 2018;125(3):790–798.

34. Wheeler MJ, Dunstan DW, Ellis KA, et al. Effect of Morning Exercise With or Without Breaks in Prolonged Sitting on Blood Pressure in Older Overweight/Obese Adults. Hypertension. 2019;73(4):859–867.

35. Duck-Won O. Community Ambulation: Clinical Criteria for Therapists’ Reasoning and Decision-making in Stroke Rehabilitation. Int J Phys Med Rehabil. 2013.

36. Carvalho LB, Kramer S, Borschmann K, Chambers B, Thijs V, Bernhardt J. Cerebral haemodynamics with head position changes post-ischaemic stroke: A systematic review and meta-analysis. J Cereb Blood Flow Metab. 2020:271678X20922457.

37. Treger I, Aidinof L, Lutsky L, Kalichman L. Mean flow velocity in the middle cerebral artery is associated with rehabilitation success in ischemic stroke patients. Arch Phys Med Rehabil. 2010;91(11):1737–1740.

38. Wheeler MJ, Dunstan DW, Smith B, et al. Morning exercise mitigates the impact of prolonged sitting on cerebral blood flow in older adults. J Appl Physiol (1985). 2019;126(4):1049–1055.

39. Biswas A, Oh PI, Faulkner GE, et al. Sedentary time and its association with risk for disease incidence, mortality, and hospitalization in adults: a systematic review and meta-analysis. Ann Intern Med. 2015;162(2):123–132.

40. Chrismas BCR, Taylor L, Cherif A, Sayegh S, Bailey DP. Breaking up prolonged sitting with moderate-intensity walking improves attention and executive function in Qatari females. PLoS One. 2019;14(7):e0219565.

41. Schreiber SJ, Gottschalk S, Weih M, Villringer A, Valdueza JM. Assessment of blood flow velocity and diameter of the middle cerebral artery during the acetazolamide provocation test by use of transcranial Doppler sonography and MR imaging. AJNR Am J Neuroradiol. 2000;21(7):1207–1211.

42. Serrador JM, Picot PA, Rutt BK, Shoemaker JK, Bondar RL. MRI measures of middle cerebral artery diameter in conscious humans during simulated orthostasis. Stroke. 2000;31(7):1672–1678.

43. Giller CA, Bowman G, Dyer H, Mootz L, Krippner W. Cerebral arterial diameters during changes in blood pressure and carbon dioxide during craniotomy. Neurosurgery. 1993;32(5):737-741; discussion 741-732.

44. Steventon JJ, Hansen AB, Whittaker JR, et al. Cerebrovascular Function in the Large Arteries Is Maintained Following Moderate Intensity Exercise. Front Physiol. 2018;9:1657.

45. Mijajlovic MD, Pavlovic A, Brainin M, et al. Post-stroke dementia - a comprehensive review. BMC Med. 2017;15(1):11.

46. Liu J, Wang Y, Akamatsu Y, et al. Vascular remodeling after ischemic stroke: mechanisms and therapeutic potentials. Prog Neurobiol. 2014;115:138–156.

47. Treger I, Luzki L, Gil M, Ring H. Transcranial Doppler monitoring during language tasks in stroke patients with aphasia. Disabil Rehabil. 2007;29(15):1177–1183.

